# Younger age of initiation of selling sex and depressive symptoms among female sex workers in Eswatini

**DOI:** 10.1101/2022.08.22.22279088

**Authors:** Ashley Grosso, Rebecca Fielding-Miller, Sindy Matse, Bhekie Sithole, Stefan Baral

**Author notes:** **Correspondence:** Ashley Grosso.

## Abstract

**Background:** Minors who sell sex are likely to have complex mental health needs that may persist into adulthood. This topic is understudied in Sub-Saharan Africa. This study hypothesized that adult female sex workers in Eswatini who started selling sex as minors have a higher prevalence of depression than those who started as adults. We also examined correlates of depression and underage initiation of selling sex, including stigma and condom-related behaviors.

**Methods:** From October-December 2014, women aged 18 or older who sold sex in the past 12 months in Eswatini were recruited through venue-based sampling. Participants completed a survey including the 9-item Patient Health Questionnaire (PHQ-9) and a question about the age at which they first sold sex for money. T-tests, χ2 tests and multivariable logistic regression were used to assess associations.

**Results:** Overall, 43.1% of participants (332/770) had probable depression, and 16.6% (128/770) started selling sex as minors under the age of 18. Over half (55.5%, 71/128) of those who started selling sex as minors had depression. This was significantly higher than the 40.7% (261/642) prevalence of depression among participants who started selling sex as adults (p=0.002). After adjusting for confounders, female sex workers who started selling sex as minors had higher odds of depression than those who started as adults (adjusted odds ratio 1.70, 95% confidence interval 1.11-2.60). Both depression and underage initiation of selling sex were associated with anticipating stigma toward sex workers in healthcare settings and the number of times the participant reported a condom slipped off or broke in the past month.

**Conclusion:** Results highlight the need for trauma-informed and adolescent-friendly mental health services in settings free of stigma toward female sex workers in Eswatini.

## 1 Introduction

Intersectional stigma and depression both commonly affect cisgender female sex workers (FSW) globally.^1,2^ The pooled prevalence of depression among FSW from low and middle income countries is 41.8%.^3^ Adolescents under the age of 18 who sell sex are considered trafficked or commercially sexually exploited children and may be particularly vulnerable to immediate and long-term mental health problems, including depression.^4^ Many FSW across Sub-Saharan Africa started selling sex while underage and experience more negative sexual health outcomes compared to FSW who started as adults.^5-8^ However, evidence on the relationship between mental health and underage initiation of selling sex is mixed. In Malawi, FSW who started selling sex while underage were 2.3 times more likely to have probable depression (15%, 5/33 v. 6%, 10/167) compared to those who started as adults, but this was not statistically significant.^9^ In Ethiopia, over one third (37.5%, 144/384) of FSW who started selling sex as minors had mild to severe depression, which was slightly higher compared to those who started as adults but not statistically significant (35.9%, 147/409).^10,11^ In contrast, in South Africa FSW who started selling sex at younger ages had lower odds of depression or PTSD.^12^ Research findings on these topics among FSW in other geographic regions are also conflicting. In the United States, FSW who started selling sex as minors were had higher odds of ever attempting suicide than those who started as adults.^13^ However in Bangladesh, FSW with any mental disorder started selling sex at significantly older ages than those without a mental disorder.^14^ The researchers surmised that younger FSW may be more resilient than older FSW.

In prior research, both depression and underage initiation of selling sex have been found to be associated with socioeconomic and demographic variables including income,^15,16^ food insecurity,^17-19^ and orphanhood.^20,21^ Depression and selling sex while underage have both been linked to healthcare issues such as perceiving or experiencing sex work-related stigma in healthcare settings^17,22^ and not being tested for HIV or aware of one’s HIV status.^6,23^ Behavioral risk factors, including not carrying condoms^5,24^ and reporting condom use errors such as slipping and breaking^25,26^ are related to both depression and underage initiation of selling sex. Both FSW with depression and FSW who started selling sex as minors report a longer length of time in the sex trade^8,27^ and greater frequency of selling sex^28,29^ compared to FSW who are not depressed and FSW who started selling sex as adults.

There remains limited research about mental health in Eswatini (formerly Swaziland), a country in southern Africa. One psychiatric hospital and one psychiatrist are available to serve the population of million people.^30,31^ The prevalence of depressive disorders in Eswatini is 4%.^32^ Over one quarter of people living with HIV in Eswatini are estimated to have depressive symptoms.^33^ In a nationally representative study in Eswatini, one in ten reported suicidal ideation, plans and/or attempts in the past 12 months.^34,35^ The prevalence of suicidal ideation in the past 12 months is 17% among students aged 12-17^36^ but 42% among sexually active female students in that age group.^37^ Research has suggested that youth unemployment in Eswatini increases mental health problems and leads to selling sex.^38^ Sex work is criminalized in Eswatini.^39^ In a 2011 study, 59% of FSW in Eswatini ever had suicidal ideation, and 63% felt sad or had a depressed mood for more than two weeks at a time in the last three years.^40^ HIV prevalence among FSW was 60%, compared to about 25% among the adult population overall.^41^ About one quarter of FSW reported that they started selling sex as minors.^42^ The present study examined the relationship between depression and underage initiation of selling sex among FSW in Eswatini.

## 2 Methods

### 2.1 Participants

FSW who exchanged sex for money, favors, or goods in the past 12 months and who were at least 18 years old, assigned female sex at birth, and provided written informed consent in English or SiSwati were eligible to participate.^43,44^ 781 FSW participants completed an in-person interviewer-administered survey. For these analyses, the sample was restricted to 770 FSW with data on age of initiation of selling sex and depressive symptoms.

### 2.2 Data collection

Data collection took place from October to December of 2014 in five towns in Eswatini: Mbabane, Manzini, Piggs Peak, Lavumisa, and Nhlangano. Participants lived in the towns and surrounding geographic areas. These sites were selected based on region, proximity to the border, population size, presence of local partner organizations, and the existence of key populations programming, health services, and other sex work “hotspots”.

A modified version of the Priorities for Local AIDS Control Efforts (PLACE) methodology was used.^45^ FSW were recruited through sampling at “hotspot” venues that were identified through key informant interviews. Study staff verified and visited the venues and approached potential participants. Venue managers and community leaders assisted in identifying and introducing FSW to the study staff. This venue-based sampling was supplemented through peer referral and snowball sampling to reach FSW who do not frequent public venues and increase the representativeness of the sample.

Upon completion of the survey, each participant was given a 50 Emalangeni [approximately 5 United States dollars] reimbursement for transportation and offered condoms and condom-compatible lubricants. The Eswatini Ministry of Health and Scientific Ethics and the Johns Hopkins School of Public Health Institutional Review Board approved the study.

### 2.3 Measures

#### 2.3.1 Depressive symptoms

Depressive symptoms in the past two weeks were measured using the nine-item Patient Health Questionnaire (PHQ-9).^46^ The PHQ-9 in this sample had a Cronbach alpha of 0.91. PHQ-9 scores of ten or higher were considered indicative of probable depression. Depression scores were categorized by severity as minimal (0-4), mild (5-9), moderate (10-14), moderately severe (15-19), and severe (20-27). Participants were asked how difficult these symptoms made it for them to do their work, take care of things at home, or get along with other people (not difficult at all, somewhat difficult, very difficult, or extremely difficult). Participants who reported any depressive symptoms were asked if they ever sought help or treatment from a healthcare provider. Participants who never sought help or treatment from a healthcare provider were asked if the reason they did not was related at all to the fact that they sell sex.

#### 2.3.2 Underage initiation of selling sex

Age of initiation of selling sex for money was dichotomized to compare those who started before the age of 18 to those who started at age 18 or older.

#### 2.3.3 Covariates

Continuous variables included the participant’s age at the time of the study, number of years selling sex, days per month selling sex, and weekly income in emalangeni and the number of times the participant reported a condom slipped off or broke in the past month.

Dichotomous variables included whether the participant reported that her parents died before she was 18 years old, started selling sex to feed herself/her family, and was ever afraid of or avoided seeking healthcare due to worrying that someone would find out she sells sex.

Categorical variables included the participant’s frequency of carrying condoms while selling sex (never, almost never, sometimes, almost always/often, or always) and the participant’s self-reported HIV status (never tested, HIV-negative, or living with HIV).

### 2.4 Analyses

Analyses were conducted using Stata/MP17.0. PHQ-9 scores by younger or older age of initiation of selling sex were compared using a t-test. χ2 tests were used to compare by age of initiation of selling sex the prevalence of depression, depression severity, each depressive symptom (i.e., any response other than “not at all”), seeking treatment, not seeking treatment because of selling sex, and difficulties due to depressive symptoms.

The relationships between each covariate and probable depression and each covariate and underage initiation of selling sex were assessed using χ2 tests for categorical variables and t-tests for continuous variables. Covariates that were significantly related to both probable depression and underage initiation of selling sex in bivariate models (p<0.05) were included in a multivariable logistic regression model of factors associated with probable depression. Missing data (<5%) were handled using listwise deletion.

## 3 Results

### 3.1 Descriptive statistics

As Table 1 shows, the mean age of participants at the time of the study was 27.5. 7.6% of participants (57/747) lost both of their parents and 34.7% (259/747) lost at least one parent before the age of 18. Most participants (81.4%, 627/770) started selling sex to feed themselves or their families. Participants had been selling sex on an average of 6.2 years and sold sex on an average of 14.8 days per month. Their average weekly income was 908.5 emalangeni, or about $62 USD. Most participants (87.4%, 673/770) always carried condoms when selling sex. The mean number of times participants reported that a condom slipped off or broke was once in the past month. About one quarter of participants (25.3%, 195/770) had ever been afraid of or avoided seeking healthcare because of worrying someone would find out they sell sex. 12.9% of participants (97/750) had never been tested for HIV, 54.0% (405/750) had tested negative, and 33.1% (248/750) had tested positive. 16.6% of participants (128/770) started selling sex as minors. 43.1% of participants (332/770) had probable depression.

**Table 1.** Characteristics of female sex workers in Eswatini by probable depression and age of entry into selling sex, 2014

|  | Total | Not depressed | Depressed | P value | Started selling sex 18+ | Started selling sex <18 | P value |
| --- | --- | --- | --- | --- | --- | --- | --- |
| Depressed | 43.1%<br>(332/770) | - | - | - | 40.7%<br>(261/642) | 55.5%<br>(71/128) | 0.002 |
| Started selling sex <18 | 16.6%<br>(128/770) | 13.0%<br>(57/438) | 21.4%<br>(71/332) | 0.002 | - | - | - |
| Mean current age | 27.5 | 27.5 | 27.5 | 0.868 | 28.0 | 24.8 | <0.001 |
| Orphaned <18 |  |  |  |  |  |  |  |
| 2 parents died | 7.6%<br>(57/747) | 8.0%<br>(34/427) | 7.2%<br>(23/320) | 0.693 | 5.8%<br>(36/624) | 17.1%<br>(21/123) | <0.001 |
| 1+ parents died | 34.7%<br>(259/747) | 36.5%<br>(156/427) | 32.2%<br>(103/320) | 0.217 | 30.9%<br>(193/624) | 53.7%<br>(66/123) | <0.001 |
| Started selling sex to feed self/family | 81.4%<br>(627/770) | 77.2%<br>(338/438) | 87.1%<br>(289/332) | <0.001 | 82.7%<br>(531/642) | 75.0%<br>(96/128) | 0.041 |
| Mean years selling sex | 6.2<br>(n=757) | 5.9<br>(n=432) | 6.6<br>(n=325) | 0.048 | 5.8<br>(n=630) | 8.0<br>(n=127) | <0.001 |
| Mean days per month selling sex | 14.8<br>(n=767) | 14.3<br>(n=436) | 15.6<br>(n=331) | 0.017 | 14.5<br>(n=639) | 16.2<br>(n=128) | 0.026 |
| Mean weekly income in emalangeneni | 908.5<br>(n=747) | 774.4<br>(n=419) | 1079.8<br>(n=328) | <0.001 | 885.6<br>(n=622) | 1022.0<br>(n=125) | 0.110 |
| Frequency of carrying condoms when selling sex |  |  |  |  |  |  |  |
| Never | 2.9%<br>(22/770) | 1.8%<br>(8/438) | 4.2%<br>(14/332) | 0.333 | 2.0%<br>(13/642) | 7.0%<br>(9/128) | 0.023 |
| Almost never | 0.3%<br>(2/770) | 0.2%<br>(1/438) | 0.3%<br>(1/332) |  | 0.2%<br>(1/642) | 0.8%<br>(1/128) |  |
| Sometimes | 4.0%<br>(31/770) | 4.6%<br>(20/438) | 3.3%<br>(11/332) |  | 4.1%<br>(26/642) | 3.9%<br>(5/128) |  |
| Almost always/often | 5.5%<br>(42/770) | 5.5%<br>(24/438) | 5.4%<br>(18/332) |  | 5.5%<br>(35/642) | 5.5%<br>(7/128) |  |
| Always | 87.4%<br>(673/770) | 87.9%<br>(385/438) | 86.8%<br>(288/332) |  | 88.3%<br>(567/642) | 82.8%<br>(106/128) |  |
| Mean times condom slipped off or broke in the past month | 1.0<br>(n=753) | 0.8<br>(n=427) | 1.3<br>(n=326) | <0.001 | 0.9<br>(n=628) | 1.3<br>(n=125) | 0.020 |
| Ever afraid of/avoided seeking healthcare because of selling sex | 25.3%<br>(195/770) | 15.8%<br>(69/438) | 38.0%<br>(126/332) | <0.001 | 23.5%<br>(151/642) | 34.4%<br>(44/128) | 0.010 |
| Self-reported HIV status |  |  |  |  |  |  |  |
| Never tested | 12.9%<br>(97/750) | 7.3%<br>(31/425) | 20.3%<br>(66/325) | <0.001 | 12.6%<br>(79/628) | 14.8%<br>(18/122) | 0.695 |
| Tested negative | 54.0%<br>(405/750) | 55.8%<br>(237/425) | 51.7%<br>(168/325) |  | 53.8%<br>(338/628) | 54.9%<br>(67/122) |  |
| Tested positive | 33.1%<br>(248/750) | 36.9%<br>(157/425) | 28.0%<br>(91/325) |  | 33.6%<br>(211/628) | 30.3%<br>(37/122) |  |

### 3.2 Relationship between selling sex while underage and depression symptoms

Over half (55.5%, 71/128) of FSW who started selling sex as minors had probable depression. This was significantly higher than the 40.7% (261/642) prevalence of depression among FSW who started as adults (p=0.002). Over one in five (21.4%, 71/332) FSW who were depressed started selling sex as minors, compared to 13.0% (57/438) of FSW who were not depressed.

As Table 2 shows, the mean PHQ-9 score was 9.18. The mean PHQ-9 score was significantly higher among FSW who started selling sex as minors compared to FSW who started as adults (10.45 v. 8.93, p=0.020).

**Table 2.** Average PHQ-9 score by age of entry into selling sex among female sex workers in Eswatini, 2014

|  | PHQ-9 score |  |
| --- | --- | --- |
|  | Median | Mean |
| Started selling sex <18 | 11 | 10.45 |
| Started selling sex 18+ | 8.5 | 8.93 |
| Total | 9 | 9.18 |
| N=770; p=0.020; PHQ-9=Patient Health Questionnaire |  |  |

As Figure 1 shows, a higher proportion of FSW who started selling sex as minors had moderate, moderately severe, or severe depression symptoms, while a higher proportion of FSW who started as adults had minimal or mild symptoms (p=0.032).

**Figure 1.**
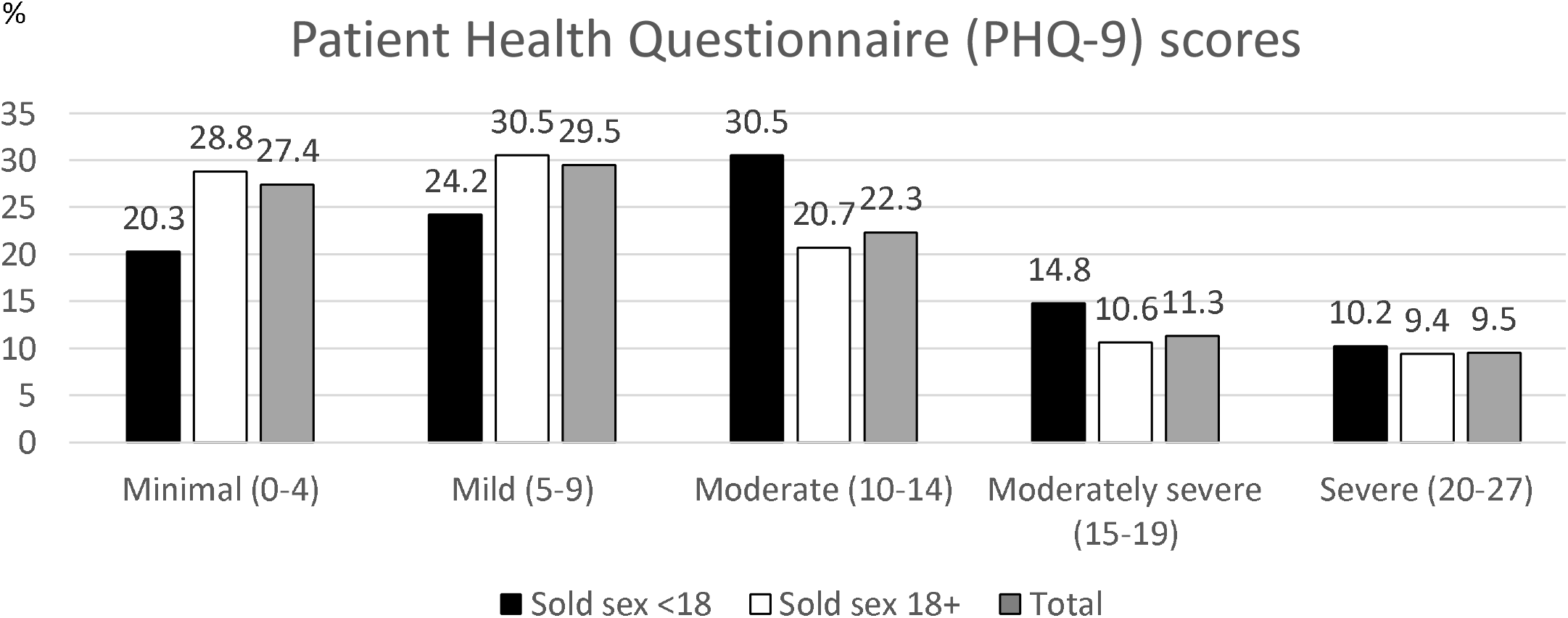
Depression severity by younger or older initiation of selling sex among female sex worker study participants in Eswatini, 2014

Table 3 shows the proportion of FSW who reported each depression symptom (i.e., any response other than “not at all”). The most common symptom was feeling down, sad, or hopeless (74.6%, 574/770). The least common symptom was suicidal ideation. This was still highly prevalent; 42.7% (329/770) of FSW had thoughts that they would be better off dead or hurting themselves in some way. A significantly higher proportion of FSW who sold sex as minors compared to FSW who started as adults reported experiencing each depression symptom except suicidal ideation and feeling tired or having little energy.

**Table 3.**
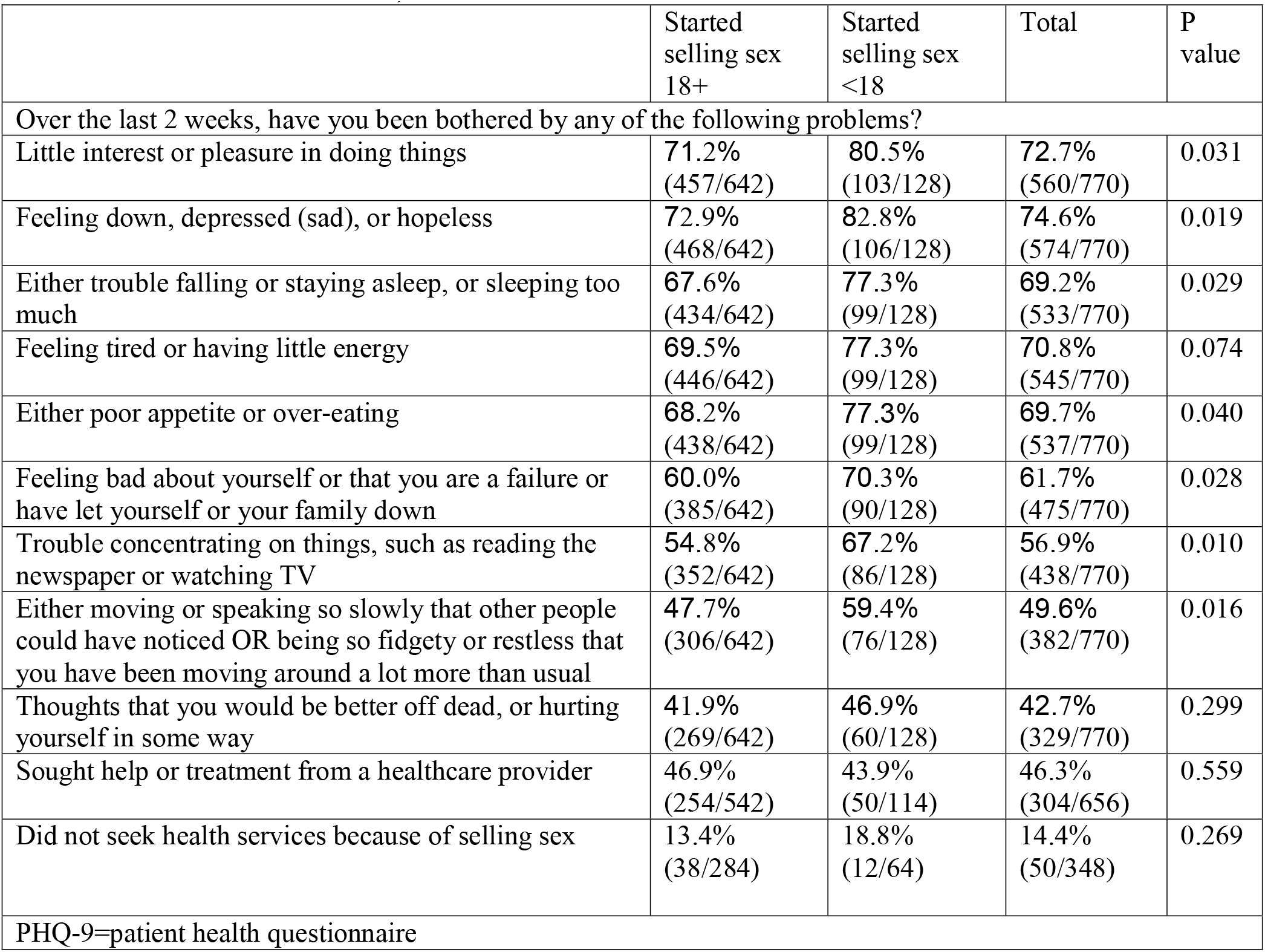
Differences in PHQ-9 items and seeking treatment by age of entry into selling sex among female sex workers in Eswatini, 2014

Less than half of those with depression symptoms (46.3%, 304/656) had sought help or treatment from a healthcare provider. This did not differ significantly based on age of initiation of selling sex (p=0.559). A small proportion (14.4%, 50/348) did not seek treatment due to the fact that they sell sex. This did not differ significantly based on age of initiation of selling sex (p=0.269).

Of those who reported any depressive symptoms, 19.1% (125/656) said these problems made it not difficult at all to do their work, take care of things at home, or get along with other people. 52.6% (345/656) said it was somewhat difficult, 21.3% (140/656) said it was very difficult, and 7.0% (46/656) said it was extremely difficult (Figure 2). This did not significantly vary based on age of initiation of selling sex (p=0.295).

**Figure 2.**
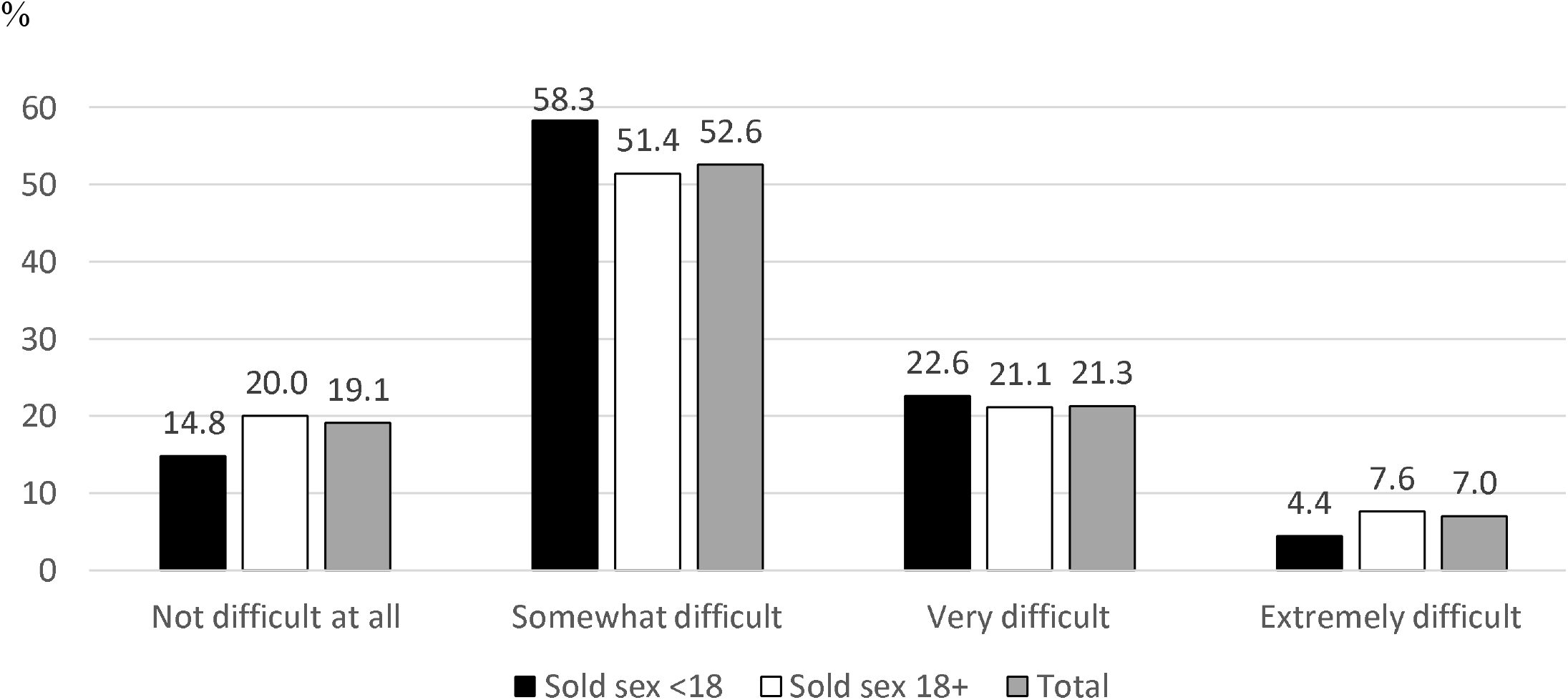
If you are having any of the problems above, how difficult have these problems made it for you to do your work, take care of things at home, or get along with other people?

### 3.3 Covariates associated with probable depression

As Table 1 shows, a higher proportion of FSW who were depressed started selling sex to feed themselves or their families compared to FSW who were not depressed (87.1% v. 77.2%, p<0.001). FSW who were depressed had been selling sex for longer (6.6 years v. 5.9 years, p=0.048) and sold sex for more days per month (15.6 v. 14.3, p=0.017) than FSW who were not depressed. The average income of FSW who were depressed was higher than that of FSW who were not depressed (1079.8 v. 774.4 emalangeni, p<0.001). Compared to FSW who were not depressed, FSW who were depressed reported that a condom slipped off or broke more often in the past month (1.3 v. 0.8 times, p<0.001). 38.0% of FSW who were depressed had ever been afraid of or avoided seeking healthcare due to selling sex, while only 15.8% of FSW who were not depressed reported this (p<0.001). One fifth (20.3%) of FSW who were depressed had never been tested for HIV, while less than one in thirteen (7.3%) FSW who were not depressed had never been tested (p<0.001). Depression was not significantly associated with current age, loss of parents, or frequency of carrying condoms.

### 3.4 Covariates associated with underage initiation of selling sex

As Table 1 shows, FSW who started selling sex as minors were younger at the time of the study than FSW who started as adults (24.8 v. 28.0, p<0.001). A higher proportion of FSW who started selling sex as minors had lost at least one (53.7% v. 3.9%, p<0.001) or both parents (17.1% v. 5.8%, p<0.001) before the age of 18 than FSW who started as adults. Three quarters (75.0%) of FSW who started selling sex as minors had done so to feed themselves or their families, while 82.7% of FSW who started as adults reported this reason (p=0.041). FSW who started selling sex as minors had been selling sex for longer (8.0 years v. 5.8 years, p<0.001) and sold sex for more days per month (16.2 v. 14.5, p=0.026) than FSW who started as adults. Compared to FSW who started selling sex as adults, a higher proportion of FSW who started as minors never carried condoms when selling sex (7.0% v. 2.0%, p=0.023). The average number of times a condom slipped off or broke in the past month was higher among FSW who started selling sex as minors than among those who started as adults (1.3 v. 0.9, p=0.020). Over one third (34.4%) of FSW who started selling sex as minors were ever afraid of or avoided seeking healthcare due to selling sex compared to less than one quarter (23.5%) of FSW who started as adults (p=0.010). Underage initiation of selling sex was not significantly associated with income or self-reported HIV status.

### 3.5 Multivariable results

The categorical covariates in the multivariable model that were significantly related to both underage initiation of selling sex and depression were whether the participant started selling sex to feed herself or her family and whether she was afraid of or avoided seeking healthcare due to selling sex. The continuous covariates in the multivariate model were the participants’ number of years selling sex, days per month selling sex, and times condoms slipped or broke in the past month (Table 4). In the multivariable model, FSW who started selling sex as minors had higher odds of having probable depression (adjusted odds ratio=1.70, 95% confidence interval=1.11-2.60). Starting sex work to feed oneself or one’s family, frequency of condoms breaking or slipping, and fear or avoidance of healthcare remained significantly associated with depression. The number of years and the number of days per month that a participant sold sex were not significantly associated with probable depression in the multivariable model.

**Table 4.** Multivariable logistic regression analysis of factors associated with probable depression among female sex worker study participants in Eswatini, 2014

|  |  |
| --- | --- |
|  | Adjusted odds ratio (95% confidence interval) |
| Started selling sex <18 | 1.70 (1.11, 2.60) |
| Started selling sex to feed self/family | 2.33 (1.51, 3.59) |
| Number of years selling sex | 1.00 (0.97, 1.04) |
| Days per month selling sex | 1.00 (0.98, 1.03) |
| Number of times a condom slipped off or broke in the last month | 1.21 (1.09, 1.35) |
| Ever afraid of or avoided seeking healthcare due to fear of someone learning she sells sex | 3.17 (2.21, 4.56) |
| N=737 |  |

## 4 Discussion

Over 40% of FSW in Eswatini in this study reported depression. These findings are consistent with other studies of FSW in sub-Saharan Africa that have shown a high burden of depression.^17,23,47-49^ In this study, 16.6% of FSW in Eswatini started selling sex as minors, which is comparable to the prevalence in other countries in the region such as South Africa (18%),^20^, Lesotho (20%),^5^ Zimbabwe (10%),^50^ and Malawi (17%).^9^ Depression and underage initiation of selling sex in this study were both associated with selling sex to feed oneself or one’s family, perceived sex work stigma in healthcare settings, and condoms breaking or slipping. Depression was also associated with income and never testing for HIV. Underage initiation of selling sex was also associated with younger age at the time of the study, being orphaned, and carrying condoms less frequently.

### 4.1 Underage initiation of selling sex and depression

This study’s findings strengthen the evidence base on the relationship between underage initiation of selling sex and depression in Africa. In Malawi, FSW who started selling sex as minors had over twice the odds of depression (aOR 2.4), but the results were not statistically significant. In our study, which had a larger sample size and higher prevalence of depression, a slightly smaller (aOR 1.70) but statistically significant difference was found. Our findings suggest the need to address the mental health needs of people selling sex in Eswatini, particularly those who are underage or started while underage. Given the limited availability of psychiatric services and high prevalence of HIV in Eswatini, a nurse counsellor-delivered brief psychological intervention that was piloted with people living with HIV, tuberculosis, and depression in Eswatini could be implemented with FSW and adolescents.^51^ Trauma-informed approaches to mental health treatment are needed for women who started selling sex as minors,^52^ while being mindful of the fact that they may still be selling sex or may not consider themselves victims of trafficking or child sexual exploitation. Preventing underage initiation of selling sex and providing alternatives to minors already selling sex to exit the sex trade through programs such as cash transfers and social protection^53^ could reduce a broad range of harms, including depression.

### 4.2 Socioeconomic implications

Consistent with research in South Africa, depression among FSW in Eswatini was associated with selling sex due to food insecurity.^17^ Scaling up existing programs that address food insecurity may have the additional benefit of decreasing depression by reducing reliance on sex work among those who do not want to sell sex.^54^ In contrast, recent depression among FSW in this study was associated with higher current income, consistent with research findings among FSW in China.^15^ Once FSW have enough income to meet their basic needs such as food, they may become more aware of their mental health needs.

Similar to research in Mexico, entry into selling sex due to food insecurity was less common among FSW in Eswatini who started selling sex as minors.^18^ Over half of FSW who started selling sex as minors had lost at least one of their parents before the age of 18, and 17.1% lost both of their parents before age 18. Those who start selling sex as minors, particularly those who were orphaned, may do so for reasons not captured in our survey, such as to pay school fees or purchase drugs or alcohol.^55^ Increasing services for orphans in Eswatini may prevent underage initiation of selling sex and subsequent negative health outcomes.

### 4.3 Healthcare service implications

Over one third of FSW who were depressed and FSW who started selling sex as minors had ever been afraid of or avoided seeking healthcare due to fear of someone learning about their involvement in selling sex. This is consistent with research in South Africa showing that depression was related to perceived or experienced sex work-related stigma in healthcare settings.^17^ Anticipating stigma toward FSW may contribute toward feelings of depression. Conversely, FSW who are depressed may have heightened fear of stigma in health settings. This fear of stigma may be related to the finding in our study that less than half of FSW with depressive symptoms sought mental health treatment, and one in eight FSW who did not seek treatment stated the reason was related to selling sex. Qualitative research with minors who sell sex in other settings has also identified stigma in healthcare as a barrier to care.^22,56,57^ Interventions to reduce stigma toward sex workers in healthcare settings that have been found to be effective in other countries could be adapted and implemented in Eswatini.

This may be particularly beneficial to improve the uptake of physical and mental health services among FSW who are depressed and those who started selling sex as minors.^58,59^ The HIV Prevention 2.0 intervention in Senegal included training healthcare workers to increase their clinical and social competence for addressing the needs of FSW. A web-based referral system provided FSW with information on where to access nonstigmatizing health services. The intervention was found to reduce perceived healthcare stigma among FSW.

Many prior studies have shown that people living with HIV have higher rates of depression.^60^ In contrast, our study showed that depression was most prevalent among those who never tested for HIV and least prevalent among those who were living with HIV. Our findings are consistent with research conducted with FSW in Uganda and Zambia showing that knowledge of one’s HIV status was associated with a decrease in depressive symptoms and research in South Africa showing that FSW who were depressed had lower odds of being tested for STIs.^17,23^ Depression could reduce the likelihood of HIV being diagnosed and treated. Addressing mental health could be a strategy to improve the HIV care continuum among FSW. Alternatively, uncertainty about their HIV status may contribute to symptoms of depression among FSW, and increasing uptake of testing may improve mental health. Integrating mental health services with HIV prevention, testing, and treatment services is warranted.

### 4.4 Behavioral program implications

FSW who were depressed and FSW who started selling sex as minors both reported that a condom had slipped or broken 1.3 times on average in the past month, potentially increasing their risk for pregnancy and HIV and STI acquisition and transmission. Access to services such as HIV and STI testing and treatment, pre- and post-exposure prophylaxis for HIV prevention, and contraception for those who do not want to become pregnant may be particularly important for these populations. This study did not include data on potential mediators of the relationship between condom breakage, depression, and underage initiation of selling sex such as substance use and condom application skills,^25^ but additional education for FSW on these topics may be necessary. Treating depressive symptoms among FSW could reduce breakage and slippage and enhance the protective benefits of condoms. However, an alternative explanation for our findings may be that FSW who are depressed or begin selling sex while underage may be more likely to notice when a condom slips or breaks.

17.2% of FSW in this study who started selling sex as minors did not always carry condoms when selling sex. This is consistent with research from Lesotho showing that underage initiation of selling sex was associated with avoiding carrying condoms due to fear of trouble with police (e.g., condoms being used as evidence to arrest them for selling sex).^5^ Further research is needed into specific barriers to carrying condoms among FSW in Eswatini. Providing condoms at sex work venues may particularly help FSW who started selling sex as minors.

### 4.5 Limitations

Data in this study were self-reported, which may lead to social desirability bias or inaccurate recall. The results from this sample of FSW recruited through venue-based sampling may not be generalizable to all FSW in Eswatini. Due to the cross-sectional nature of this study, it is not clear whether depression is the result of selling sex while underage or a result of other factors that led to underage initiation of selling sex. Longitudinal studies if ethically feasible could elucidate the timing of development of depressive symptoms among those who sell sex as minors.

### 4.6 Conclusion

This study demonstrates that depression is highly prevalent among FSW in Eswatini and associated with underage initiation of selling sex and other social and behavioral covariates. Taken together, these results indicate the need to comprehensively address intersecting risks and health conditions including early initiation of selling sex, mental health, stigma, food insecurity, and HIV risk among FSW in Eswatini.

## Data Availability

The data that support the findings of this study are available from the Eswatini Ministry of Health but restrictions apply to the availability of these data, and so are not publicly available. Data are however available from the authors upon reasonable request and with permission of the Swaziland Ministry of Health.

